# Novel Deep CNNs Explore Regions, Boundaries, and Residual Learning for COVID-19 Infection Analysis in Lung CT

**DOI:** 10.1101/2024.05.08.24307035

**Authors:** Bader Khalid Alshemaimri, Saddam Hussain Khan

## Abstract

COVID-19 poses a global health crisis, necessitating precise diagnostic methods for timely containment. However, accurately delineating COVID-19 affected regions in Lung CT scans is challenging due to contrast variations and significant texture diversity. In this regard, this study introduces a novel two-stage classification and segmentation CNN approach for COVID-19 lung radiological pattern analysis. A novel Residual-BRNet is developed to integrate boundary and regional operations with residual learning, capturing key COVID-19 radiological homogeneous regions, texture variations, and structural contrast patterns in the classification stage. Subsequently, infectious CT images undergo lesion segmentation in the second stage using the newly proposed RESeg segmentation CNN. The RESeg leverages both average and max-pooling implementations to simultaneously learn region homogeneity and boundary-related patterns. Furthermore, novel pixel attention (PA) blocks are integrated into RESeg to effectively address mildly infected regions. The evaluation of the proposed Residual-BRNet CNN demonstrates promising performance metrics, achieving an accuracy of 97.97%, F1-score of 98.01%, sensitivity of 98.42%, and MCC of 96.81%. Meanwhile, PA-RESeg achieves optimal segmentation performance with an IoU score of 98.43% and a Dice Similarity score of 95.96% of the lesion region. These findings highlight the potential of the proposed diagnosis framework to assist radiologists in identifying and analyzing COVID-19 affected lung regions. The CAD GUI diagnosis tool is provided at https://github.com/PRLAB21/COVID-19-Diagnostic-System.

## 1 Introduction

COVID-19, emerging from Wuhan, China, was swiftly disseminated worldwide in early 2020 [1], continuing to impact continents worldwide [2]. Currently, global COVID-19 cases stand at around 705 and 7 million suspects and demises, respectively. The vast majority, about 99.6%, experience mild symptoms, while 0.4% develop severe or critical conditions [3]. Common symptoms include flu-like cough, fever, fatigue, etc. However, severe cases may lead to respiratory inflammation, and alveolar and lung damage, potentially resulting in death [4]. COVID-19 pneumonia often presents with signs like pleural-effusion, ground-glass-opacities (GGO), and consolidation [5].

Typical diagnostic approaches for COVID-19 patients include gene sequencing, RT-PCR, as well as X-ray and CT imaging [6], [7]. RT-PCR is considered the standard test, but its cost limits accessibility, particularly in developing countries lacking sequencing facilities. Although RT-PCR is accurate, it typically requires up to 2 days for results and is susceptible to viral RNA instability, resulting in a detection rate of approximately 30% to 60% and requiring serial testing to mitigate false negatives [8]. Therefore, additional precise detection methods are essential for timely treatment and halting the transmission of COVID-19 infections [9].

CT imaging, available and cost-effective in hospitals, is a reliable tool for detecting, prognosing, and monitoring COVID-19 patients [10]. Common radiographic features of COVID-19 patients comprise GGO, consolidation, peripheral lung distribution, etc. [12]. Analyzing numerous CT images strains radiologists, especially in areas lacking expertise, impacting their effectiveness. Studies confirm the diagnostic accuracy in identifying lung abnormalities in COVID-19 cases, even without typical clinical symptoms, and in cases of false-negative RT-PCR results [11].

During public health crises like epidemics and pandemics, radiologists and healthcare facilities are overwhelmed. The radiologists struggle with identifying COVID-19 infection through CT scans, emphasizing the need for automated tools to improve performance and handle patient loads [13]. Prior to the current pandemic, deep learning (DL)-based systems supported radiologists in spotting lung anomalies, ensuring reproducibility, and detecting subtle irregularities not visible to the naked eye [14]. Amid the ongoing COVID-19 crisis, many research teams concentrate on creating automated systems for identifying infected individuals using CT images [15].

The unique radiographic patterns associated with COVID-19, such as region homogeneity, texture variation, and characteristic features like GGO, pleural effusion, and consolidation, have been extensively documented [16]. In this regard, we have proposed an integrated CNN framework for COVID-19 infection radiological pattern detection and analysis in CT images. Evaluation of the framework’s performance is conducted on a standard CT dataset, with efficacy comparisons made against established CNNs. Key contributions of the study encompass:

- A new two-stage framework is developed for the identification and analyzing COVID-19 infection regions in CT that integrates Residual-BRNet classification and PA-RESeg segmentation CNNs.
- A deep Residual-BRNet classifier integrates regional, edge operations, and residual learning to extract diverse features capturing COVID-19 radiological homogeneous areas, texture variations, and boundary patterns. Moreover, residual learning is implemented to reduce the chance of vanishing gradient.
- A newly introduced RESeg CNN accurately identifies COVID-19 affected areas within the lungs. This model systematically incorporates both average- and max-pooling implementation across encoder and decoder blocks to leverage region homogeneity and inter-class/heterogeneous features.
- The inclusion of a novel pixel attention (PA) block within RESeg effectively mitigates sparse representation issues, leading to improved segmentation of mildly infectious regions. Finally, the proposed detection and segmentation techniques are fine-tuned through TL and assessed against existing techniques.

The paper follows this structure: Section 2 offers an overview of previous COVID-19 diagnosis research. Section 3 delineates the developed framework, while Section 4 elaborates on its experimental dataset and performance metrics. Section 5 assesses results and discusses experimental evaluation, and Section 6 provides conclusions.

## 2 Related Works

Currently, CT technology is employed for COVID-19 analysis globally, including in developed and under-developed countries. However, CT scan analysis is often slow, laborious, and susceptible to human error. Consequently, DL-based diagnostic tools have been developed to expedite and improve image analysis, aiding healthcare professionals [17]–[19]. DL techniques have demonstrated optimal performance in image analysis with deep CNNs being particularly prominent [20].

Several recent CNNs have been utilized to analyze COVID-19-infected CT scans, employing diverse approaches [21]. Additionally, researchers have explored Transfer Learning (TL) to predict COVID-19-infected CTs, achieving accuracies ranging from 87% to 98% [22]–[24]. For instance, COVID-Net, inspired by ResNet, achieves an accuracy of 92% in differentiating multiple types of COVID-19 infections but with a detection rate of 87% [25]. Similarly, COVID-CAPS, achieves high accuracy (98%) but lower sensitivity (80%) to COVID-19 infection [26]. Additionally, COVID-RENet, a recently developed classification model, incorporates both smooth and boundary image features, achieving a 97% accuracy rate. These models are first trained on normal data and subsequently fine-tuned with COVID-19 specific images [27]. Alternatively, segmentation is commonly utilized to determine infection location and severity. While classical methods like watersheds were initially used, they often exhibit good performance [28]. As a result, a DL-based method named ‘VB-Net’ was implemented for segmenting COVID-19 lesions in CT, achieving a quantified 91% dice similarity (DS) score [23]. Additionally, the COVID-19 JCS system integrates both detection and analysis features to visualize and delineate infected areas, achieving a 95.0% detection rate and 93.0% specificity in classification, albeit with a lower 78.30% DS score in segmentation [29]. Furthermore, the DCN method is proposed for COVID-19 analysis, showcasing 96.74% accuracy and infection analysis DS of 83.50% [30].

However, the majority of current COVID-19 analyses have been trained on limited CT datasets, facing two primary challenges: 1) insufficient data availability, necessary for ensuring the robustness of deep CNNs across diverse COVID-19 infections; 2) limited detection capabilities, focusing solely on classifying infected samples without providing information on the location and severity of the infection.

## 3 Methodology

The research presents a new CNN-based diagnosis approach for automated COVID-19 abnormality analysis in lung CT. The framework is structured into two stages: classification and segmentation. Initially, the classification model discriminates between individuals with COVID-19 infection and healthy CT samples. PA-RESeg, a Region Estimation-based segmentation CNN, has been introduced subsequently for analyzing infectious lung regions This framework introduces three key technical novelty: (1) the Residual-BRNet classification technique, (2) the PA-RESeg segmentation technique, and (3) the implementation of customized classification and analysis CNNs. Moreover, the framework conducts segmentation of the COVID-19 lesion region in CT to capture detailed region information, assisting in assessing infection spread. Figure 1 shows the developed framework, encompassing both summary and detailed processes.

**Figure 1:**
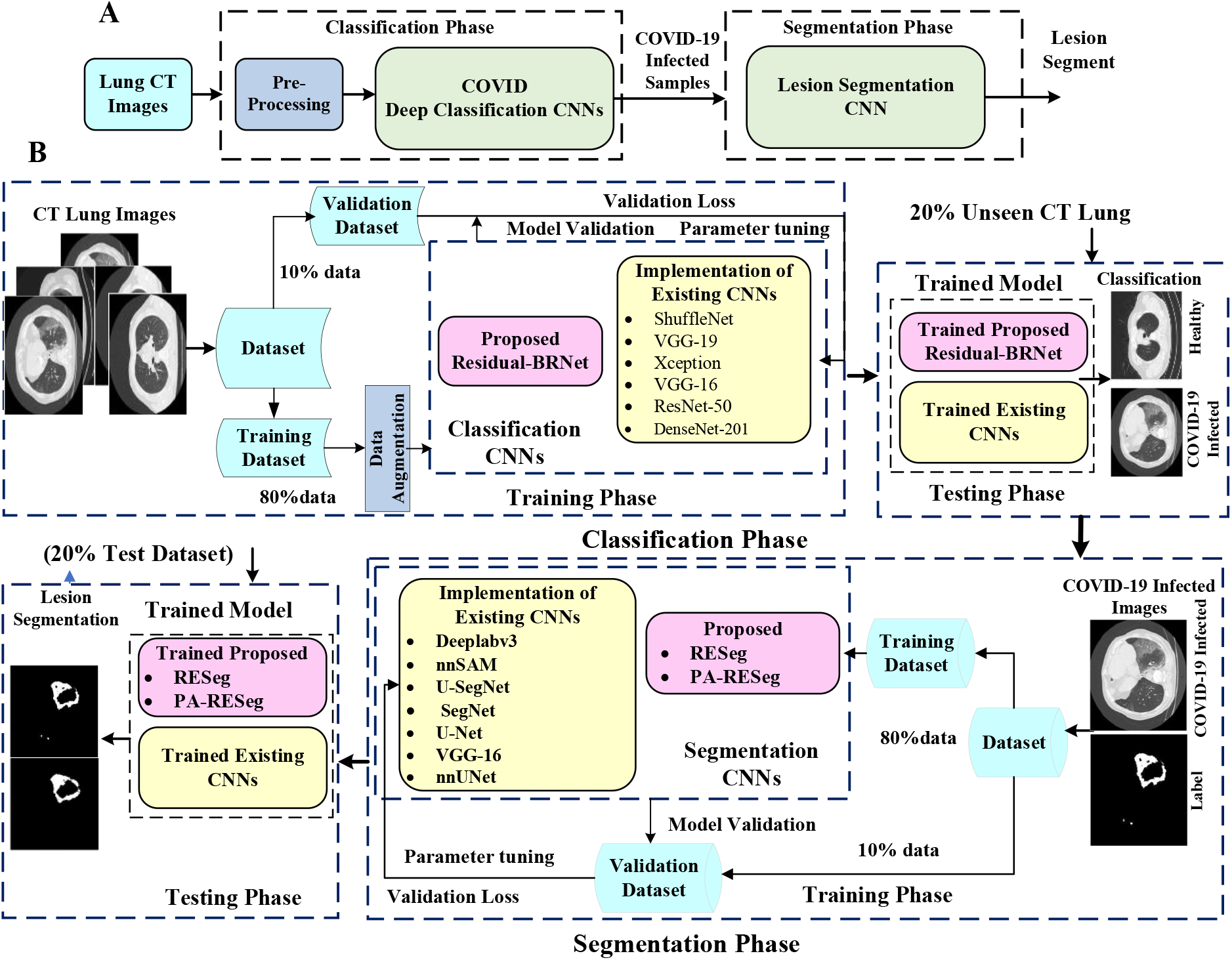
Panels A and B outline the key steps of the proposed two-stage framework and a detailed illustration of the complete workflow, respectively.

### 3.1 COVID-19 Infected CT Classification

Currently, COVID-19 infected CT samples are classified on a broad scale to differentiate them from healthy samples. Initially, in the classification stage, two distinct experimental configurations are utilized: (1) the newly developed Residual-BRNet (explained in section 3.1.1), and (2) fine-tuning deep CNN via weight transfer. Implementation setups are provided below.

#### 3.1.1 Proposed Residual-BRNet

This study presents a novel Residual-BRNet, a residual learning-based CNN, developed to distinguish COVID-19 infectious CT. The Residual-BRNet consists of four unique residual blocks comprised of distinct convolutional (Conv) based feature extractions, followed by novel homogenous, and boundary operations. These components systematically extract homogeneous region and boundary features specific to COVID-19 at each stage. Within the residual block, Conv blocks are linked with shortcut connections to optimize Conv filters and capture textural variations. Average- and max-pooling are implemented to retain relevant infected patterns, specifically homogeneous and deformed regions. The architectural layout of Residual-BRNet is illustrated in Figure 2. In each block’s conclusion, a pooling operation with a stride of two is conducted to manage model complexity and improve invariant feature learning [31].

**Figure 2:**
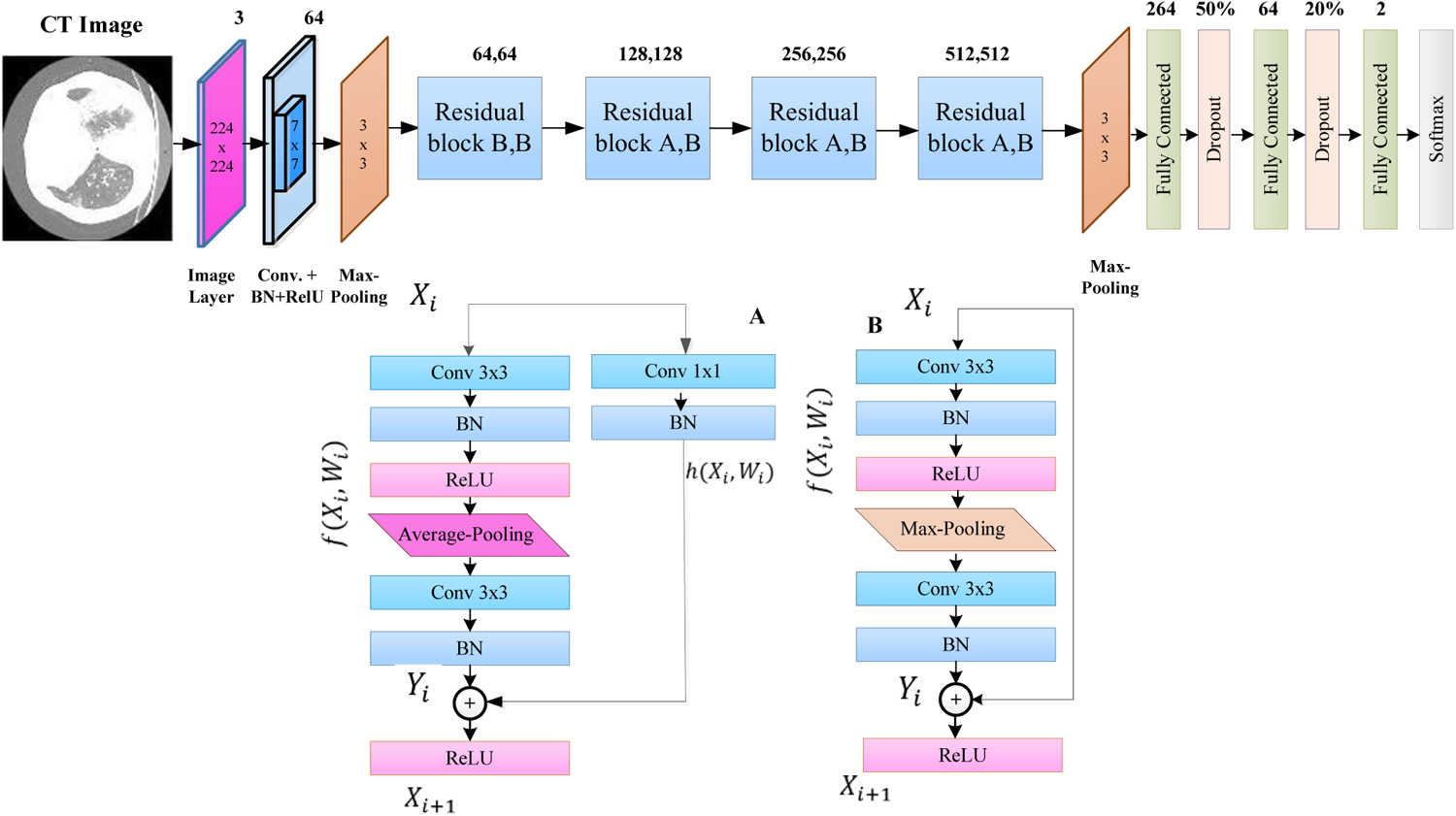
The proposed Residual-BRNet architecture.

The mathematical operations within the Conv block are detailed in Equations (1) and (3), while the residual block is outlined in Equations (4 & 5). The Conv block includes a Conv layer, BN, and ReLU for both the nth and n-1 layers. Equation (1) describes Conv operation 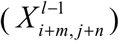 and filter 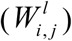 for the *l*^th^ layer. *M* × *N*, and *D* signify the resolution and depth, respectively.

Equation (2) denotes the batch-normalization (BN) for Conv outcome (*C*^*l*^), whereas *μ*_*B*_ and 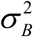 characterizes the mean and variance. Additionally, residual learning offers advantages by diminishing gradient issues, enhancing feature-map representation, and fostering convergence. Equations (3) *f* (*c*) show activation, while Equations (4 & 5) demonstrate the residual learning process, 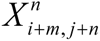 and 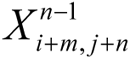 are outputs of *Block*_*n*_ and *Block*_*n*−1_, respectively. Finally, equations (6) depict the fully connected layer to reduce the feature space assessing their significance, where 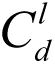 represents the Conv and *u*_*k*_ is *k*^th^ neuron. Moreover, the cross-entropy (*L*) loss activation function (presented in Equation (7)), *P*_*CT*_ represents the predicted class.

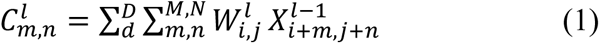

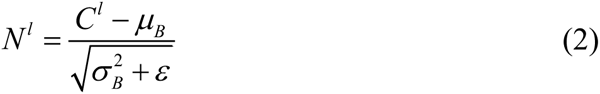

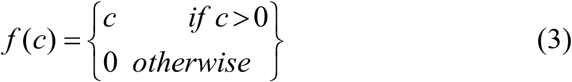

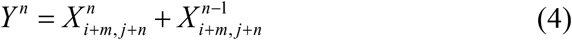

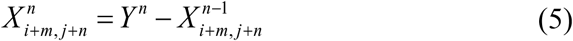

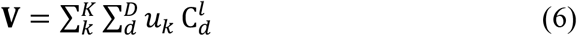

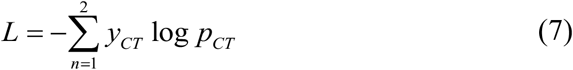

#### 3.1.2 Implementation of Existing Classification CNNs

Deep CNNs, a form of DL model, exploit spatial correlations and have demonstrated good outcomes, especially in biomedical imaging [32]–[34]. To compare the proposed Residual-BRNet, we utilized several established CNN models with variations in depth and architecture to detect COVID-19 infection in CT. The detection CNNs used comprise VGG, ResNet, DenseNet, ShuffleNet, and Xception [35]–[40]. These CNNs underwent fine-tuning using TL for comparison. Deep CNN architectures typically require a substantial dataset for efficient learning. Therefore, TL was utilized to leverage the learning from pre-trained on extensive benchmarked datasets like ImageNet [41]. In this scenario, optimizing existing deep CNN models using TL involves adapting the architecture to match CT samples and replacing final layers with the target class.

### 3.2 COVID Infection Segmentation

Accurately identifying and quantifying the infected region is important for analyzing radiological patterns and severity in diagnosis. Semantic segmentation, performed after initially distinguishing CT images at an abrasive scale, offers detailed insights into the infected areas. COVID-19 infections are separated from the surrounding areas through binary labeling pixels within the infection as the positive class, and considering all others as healthy (background). Semantic segmentation involves pixel-level classification, assigning each pixel to its respective class [42]. This study employed two distinct segmentation setups: (i) the proposed PA-RESeg, and (ii) the implementation of segmentation CNN, details are provided below.

#### 3.2.1 Proposed PA-RESeg Technique

The proposed RESeg segmentation CNN, features two encoder and decoder blocks meticulously designed to enhance feature learning. Our approach systematically integrates average with max-pooling in the encoding stages (Equations (8-9)). We employ a combination of average pooling and max-unpooling, distinguishing our model from others in the field at the decoder part. The architectural design of RESeg is depicted in Figure 3. Distinguishing between COVID-19 infectious regions and background areas presents challenges due to poorly defined borders and potential overlap with healthy lung sections. To overcome this, we use max pooling to capture boundary information, while average pooling evaluates the homogeneity of the COVID-19 infectious region.

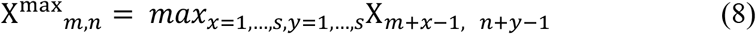

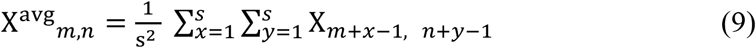

**Figure 3:**
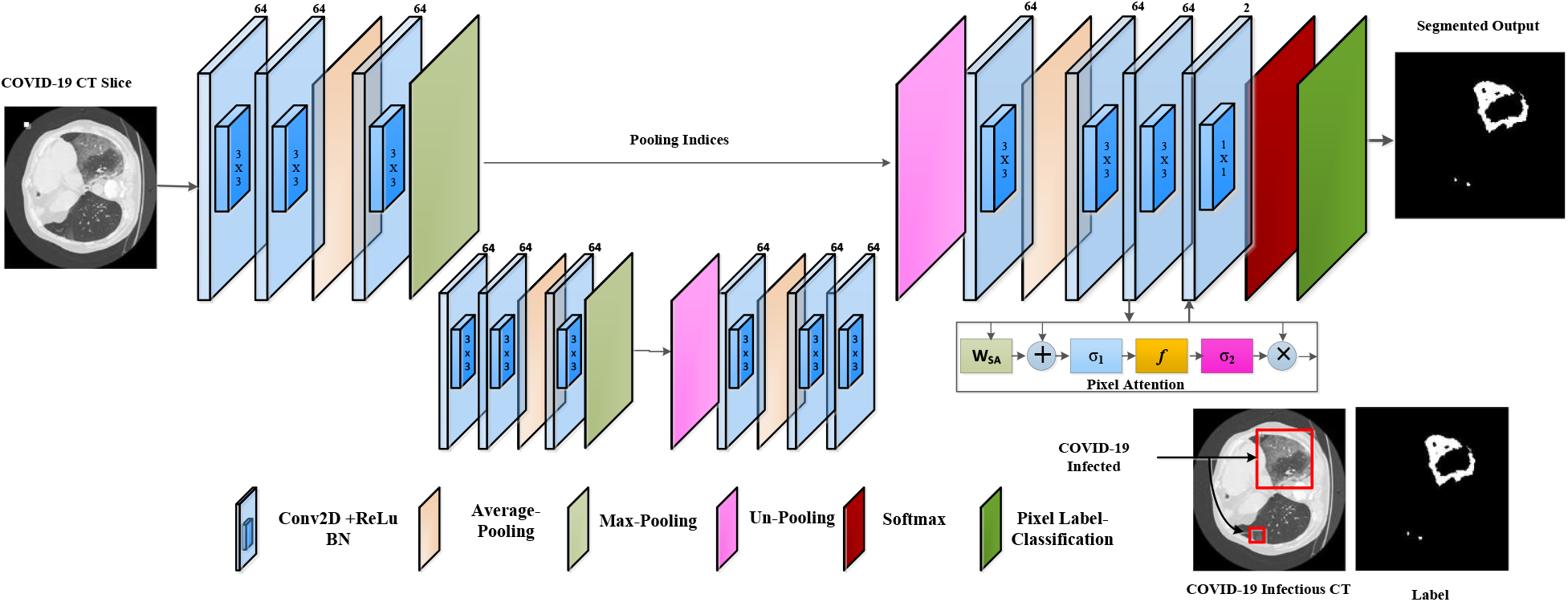
The proposed PA-RESeg architecture.

In Equations (8) and (9), **X**^avg^ and **X**^max^ as representing the average and maximum pooling operations with ‘s’ stride, respectively, applied to the convolved output (**X**_*m,n*_). We utilized an encoder-decoder technique for precise segmentation, capitalizing on the encoding stages’ capacity to learn semantically significant object-specific details. However, the encoding process can result in the loss of spatial information critical for object segmentation. To resolve this, in the decoding stage, the encoder’s channels are reconstructed using max-pooling indices to locate infectious regions. The final layer employs a 2x2 Conv operation to classify each pixel as a COVID-19 infectious region or background (healthy) using cross-entropy activation. The encoder-decoder design exhibits symmetry, with the encoder’s max-pooling layer replaced by the decoder’s un-pooling layers.

#### 3.1.1 New Pixel Attention Block

This study presents a novel technique for focusing on individual pixels during training, guided by their representation to address the unrecognized mildly infected regions [43]. This Pixel attention (PA) method emphasizes COVID-19 infection with a high weightage while assigning a lower weight to background region pixels. This strategy is incorporated into the proposed RESeg, with details depicted in Figure 4.

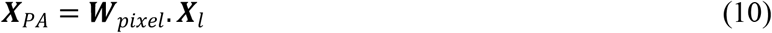

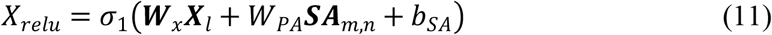

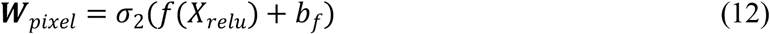

**Figure 4:**
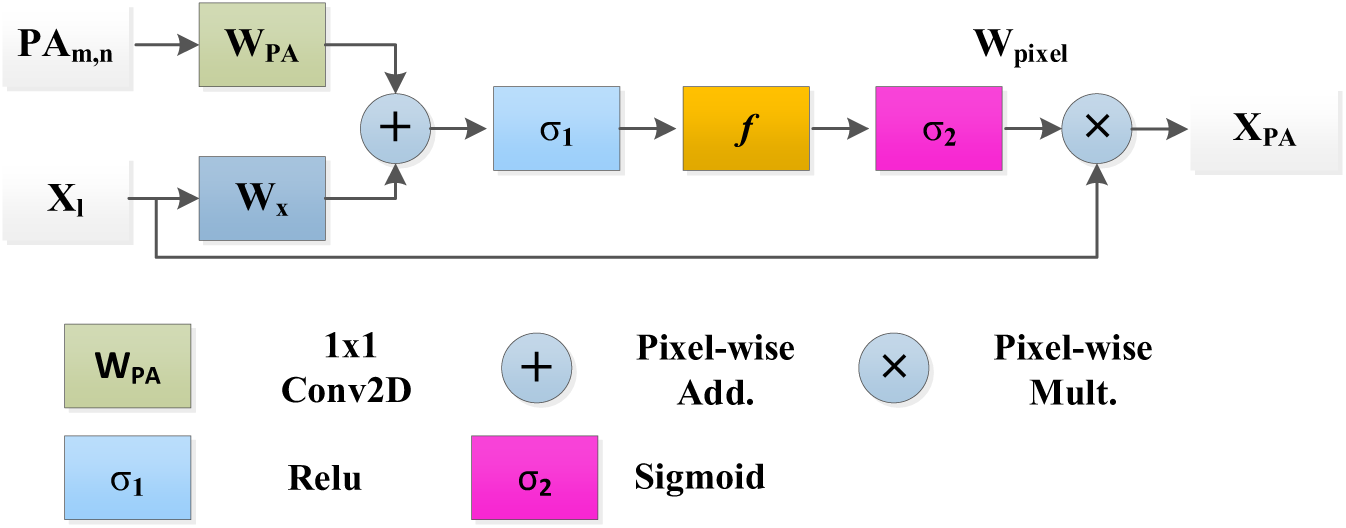
Pixel Attention Block.

Equation (10) defines ***X***_*l*_ as the input map and *W*_*pixel*_ as the pixel-weightage within the range of [0, 1]. The outcome ***X***_*PA*_ delineates the infectious area and attenuates redundant information. Equations (11) and (12) utilize *σ*_1_ and *σ*_2_ as the ReLU and Sigmoid activation, respectively. *b*_*SA*_ and *b*_*f*_ represent biases, and *W*_*x*_, *W*_*PA*_, and *f* is the linear transformation.

#### 3.1.2 Existing Segmentation CNNs

Various DL techniques with diverse architectures have been proposed and evaluated for semantic segmentation across various datasets and categories [44]. These models vary in terms of encoders and decoders, upsampling methods, and skip connections. In this study, we customized segmentation models, including nnSegment Anything Model (SAM), nnUNet, VGG-16, SegNet, U-Net, U-SegNet, and DeepLabV3 [45]–[49], for application on the COVID-19 lesion. We modified the segmentation CNN by replacing the initial and target with customized layers adjusted to the data dimensions.

## 4 Experimental Configuration

### 4.1 Dataset

The proposed diagnosis utilized a standardized CT image prepared by the Italian Society of Radiology (SIRM) and UESTC-COVID-19 Radiological Center [50]. The dataset consists of 70 patients with 10838 used axial CT samples, meticulously reviewed by experienced radiologists, with marked infected lung segments. Of these samples, 5152 display COVID-19 infection patterns, while 5686 are healthy and the whole dataset distributions are available in Table 1. Every CT sample includes a binary mask provided by a radiologist (ground truth), offering detailed pixel-level binary labels. The dataset encompasses diverse infection levels, encompassing mild, moderate, and severe. To optimize computational efficiency, all images have been dimensioned as 304x304x3 using interpolation. See Figure 5 for illustrations of COVID-19 infected and healthy images.

**Table 1.**
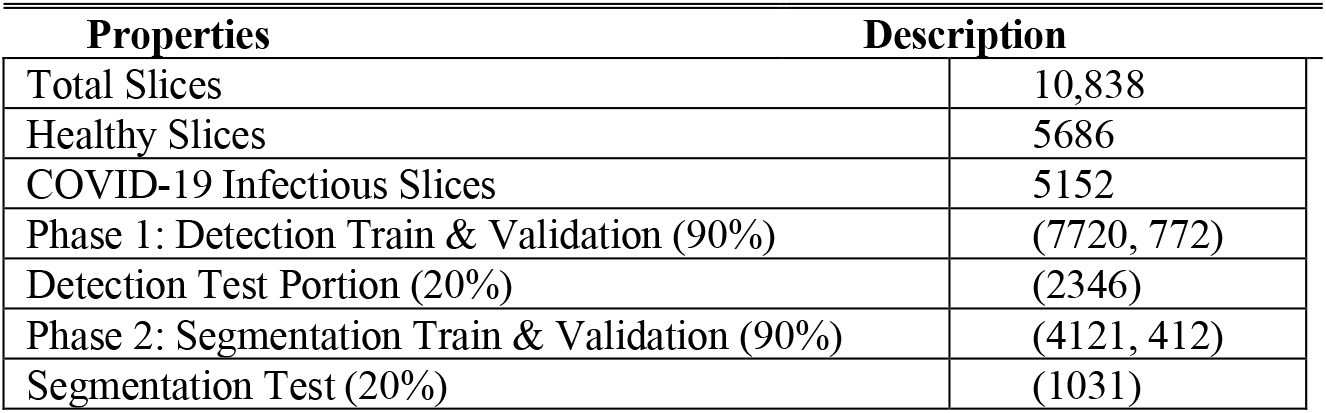
COVID-19 CT data distribution.

**Figure 5:**
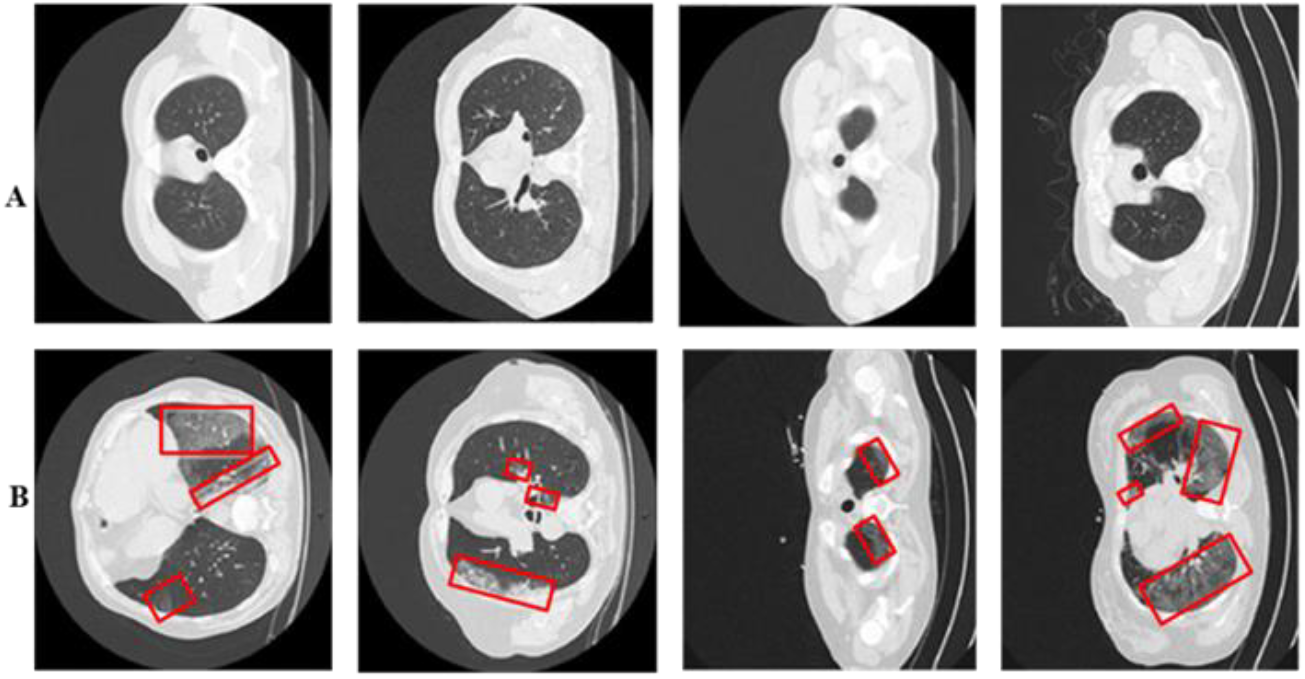
Panel (A) displays healthy lung CT samples, while Panel (B) showcases COVID-19 infected lung CT samples. Red boxes highlight regions of infection.

### 4.2 Implementation Details

The framework entails training in separate classification and analysis CNNs. The COVID-19 dataset comprises 10838 CT for detection, with 5152 infected and 5686 healthy images. These infected images and their labels are used for segmentation model training. We maintain a fixed experimental setup for both stage models, with an 8:2 split ratio for training and testing. Additionally, the training portioned into train and validation sets at a 9:1 ratio for hyperparameter selection. Cross-validation improves model robustness and generalization during hyperparameter selection. Hold-out cross-validation is employed for training both detection and segmentation CNNs. Hyperparameters are crucial for optimizing deep CNN models trained with SGD to minimize cross-entropy loss. The models undergo 30 epochs with selected optimal hyperparameters (learning rate (0.001), batch-size (8), momentum (0.95)) to ensure convergence [51]. Softmax is employed for class probability assignment in both classification and segmentation tasks. The 95% confidence interval (CI) for sensitivity and the Area Under Curve (AUC) of detection models is computed [52], [53]. MATLAB 2023b is used on an Intel Core i7 processor and Nvidia GTX 1080 Tesla GPU-enabled system. Training all networks takes approximately 3 days.

### 4.3 Performance Evaluation

The diagnosis framework’s performance is evaluated using detection and segmentation metrics. Detection metrics, including accuracy (Acc), sensitivity (S), precision (P), specificity (Sp), MCC, and F-score, are accompanied by equations (13-18). Segmentation models are assessed based on segmentation accuracy (S-Acc), IoU, and DS coefficient, presented in equations (19) and (20). Accuracy represents the accurate segregation of infected and healthy class samples, while S-Acc indicates the accurate prediction of infected and healthy pixels. The DSC metric evaluates structure similarity, and IoU quantifies the overlapping ratio between detected and label. Further explanation of performance metrics is available in Table 1.

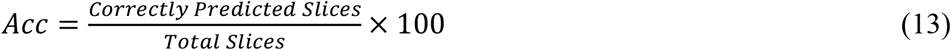

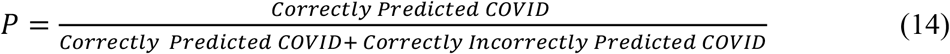

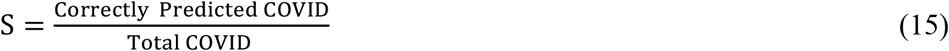

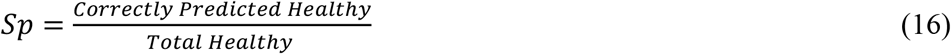

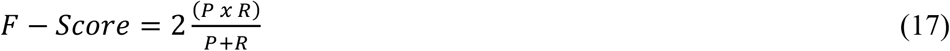

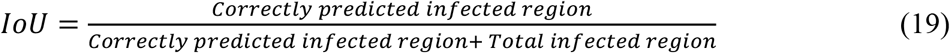

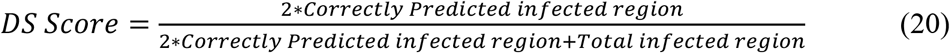

## 5 Results

This study introduces a two-stage framework for analyzing infected CT samples, followed by exploring lung infection patterns within classified COVID-19 images. This staging process reflects clinical workflows, where patients undergo additional diagnostic tests after detection.

### 5.1 COVID-19 Infected CT Classification

In this study, we introduce Residual-BRNet CNN for initial screening and categorize the infected and healthy images. The Residual-BRNet optimized performance for high detection rates of COVID-19 characteristic patterns and minimal false positives is demonstrated in Table 2. We evaluate Residual-BRNet’s learning potential for COVID-19 patterns by comparing its performance with existing CNNs.

**Table 2.** Comparison of the developed Residual-BRNet with existing CNNs.

| **CNNs** | Acc. | F-score | Pre. | MCC | Spec. | Sen. |
| --- | --- | --- | --- | --- | --- | --- |
| ShuffleNet | 89.88 | 90.00 | 88.85 | 79.76 | 88.55 | 91.38 |
| VGG-19 | 92.26 | 92.44 | 92.78 | 81.87 | 90.96 | 92.18 |
| Xception | 94.35 | 94.43 | 94.15 | 87.21 | 93.98 | 93.94 |
| VGG-16 | 95.83 | 95.81 | 97.56 | 90.67 | 96.99 | 94.67 |
| ResNet-50 | 96.13 | 96.12 | 97.58 | 91.52 | 97.59 | 95.35 |
| DenseNet-201 | 96.73 | 96.77 | 96.49 | 92.02 | 96.39 | 96.71 |
| **Proposed Residual-BRNet** | **97.97** | **98.01** | **97.61** | **96.81** | **97.62** | **98.42** |
| **Reported Studies** |  |  |  |  |  |  |
| **JCS** [29] | --- | --- | --- | --- | 93.17 | 95.13 |
| **VB-Net** [23] | --- | --- | --- | --- | 90.21 | 87.11 |
| **DCN** [30] | --- | 96.74 | --- | --- | --- | --- |
| **3DAHNet**[54] | -- | --- | --- | --- | 90.13 | 85.22 |

#### 5.1.1 Proposed Residual-BRNet Performance Analysis

The proposed Residual-BRNet ‘s performance is assessed on the test set, measuring Accuracy, F-score, MCC, sensitivity, specificity, and precision (Table 2). Compared to baseline DenseNet, Residual-BRNet exhibits superior generalization, with higher F-score (Residual-BRNet: 98.01%, DenseNet: 96.77%), accuracy (Residual-BRNet: 97.97%, DenseNet: 96.73%), and MCC (Residual-BRNet: 96.81%, DenseNet: 92.02%). The discriminative ability of Residual-BRNet is further demonstrated in the PCA plot. Additionally, a comparative feature-based analysis of the best-performing DenseNet models is presented in Figure 6 for reference.

**Figure 6:**
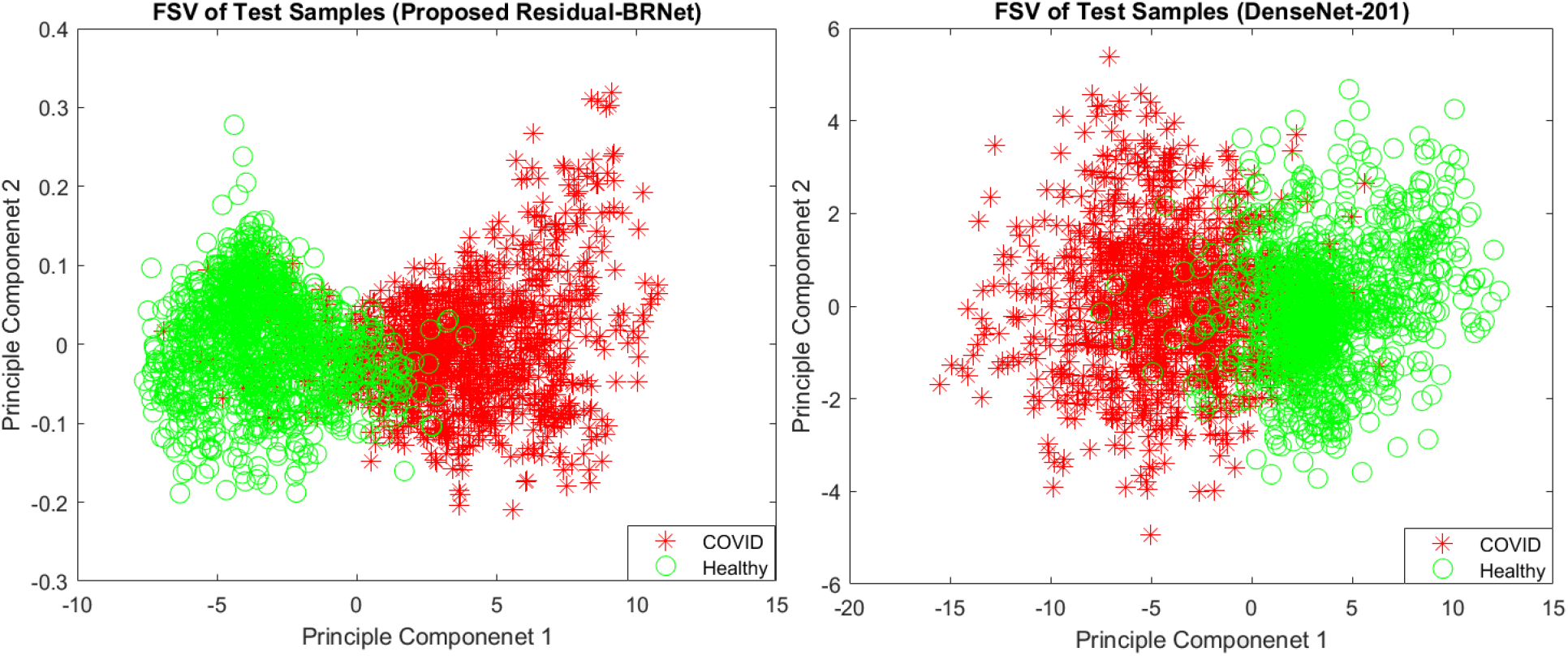
PC1/PC2-based Feature space visualization of the techniques.

#### 5.1.2 Existing CNNs Performance

The Residual-BRNet’s effectiveness is benchmarked against established deep CNN-based detection CNNs, renowned for their effectiveness in tasks such as lung abnormality classification. TL facilitates the learning of COVID-19-specific features more efficiently from CT images. Conversely, Residual-BRNet demonstrates superior performance in F-score, MCC, and accuracy (Table 3) compared to established CNNs. Notably, Residual-BRNet significantly improves classification performance by approximately (1.24 to 8.01 %) for F-score, (4.79 to 17.05%) for MCC, and (1.24 to 8.09%) for accuracy. Figure 7 illustrates the performance enhancement of Residual-BRNet over the maximum, and minimum-performing deep CNNs in terms of detection metrics.

**Table 3:**
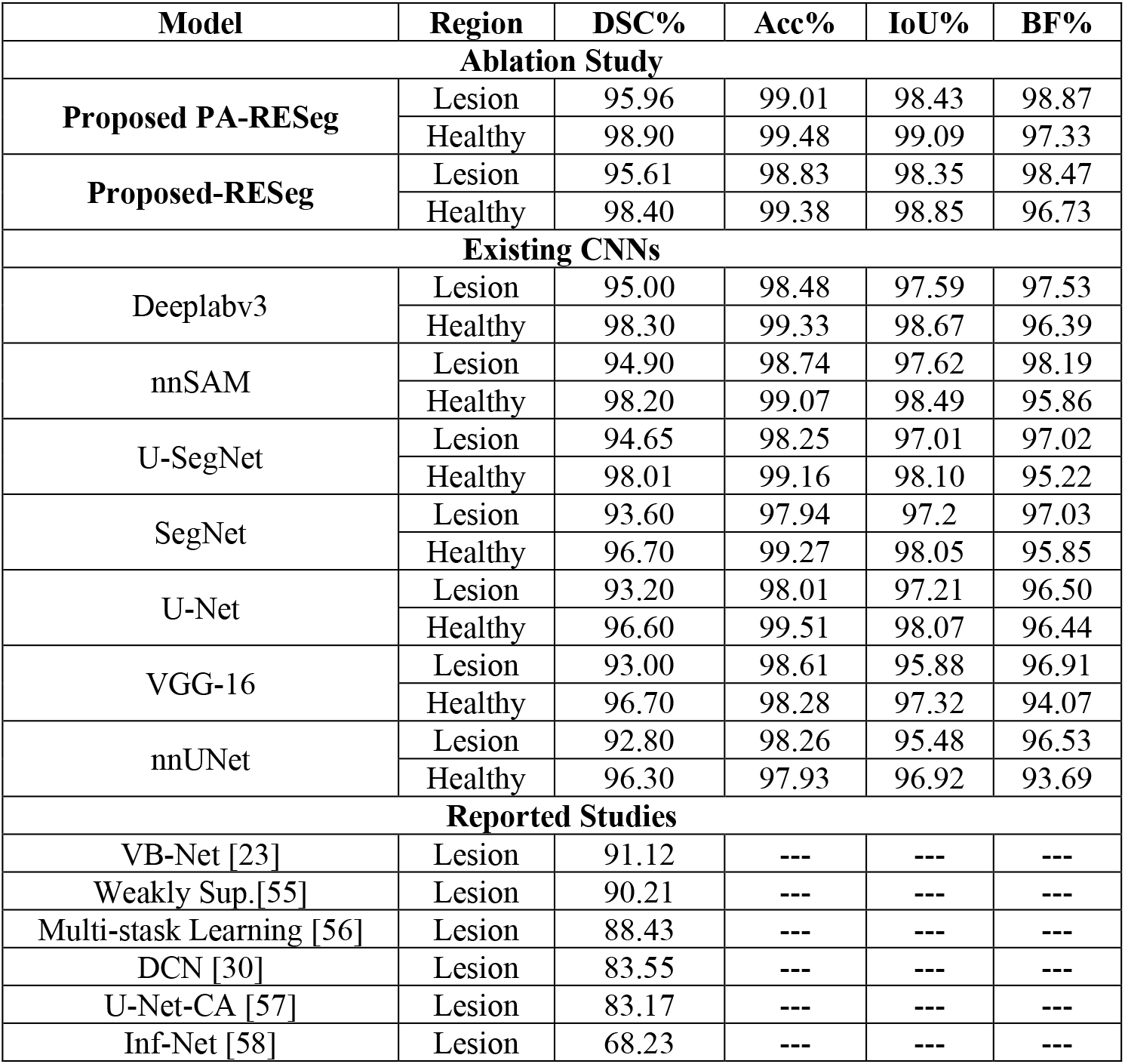
The developed PA-RESeg and current segmentation CNNs Analysis.

**Figure 7:**
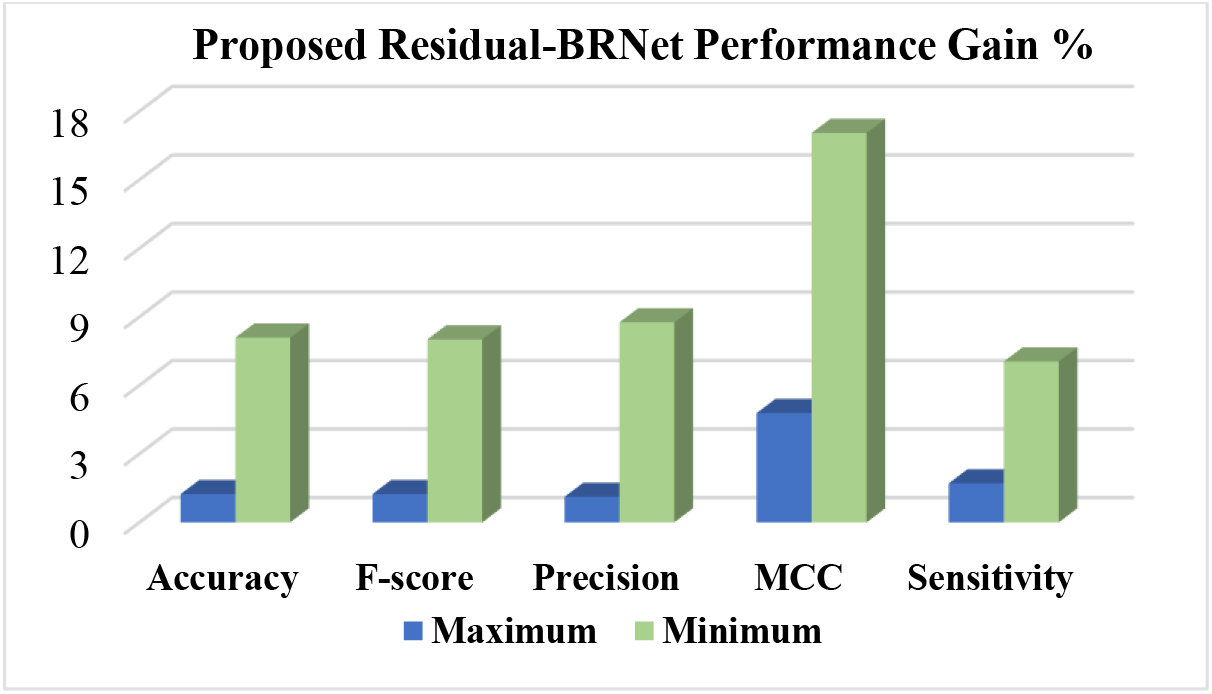
The performance evaluation of the proposed Residual-BRNet is conducted, assessing detection metrics.

#### 5.1.3 PR and ROC curve-based comparison

PR and ROC curves are utilized for quantitative evaluation of the segregation ability of detection CNNs, depicted in Figure 8. These curves act as performance metrics, assessing the classifier’s generalization ability by showcasing the distinction of inter-class variation across varying thresholds. PR curves for Residual-BRNet and existing CNN, demonstrating the superior learning capacity of the proposed CNN. The proposed Residual-BRNet outperforms DenseNet and other deep CNN models in terms of AUCs, F-score, Accuracy, and MCC overall (Table 2).

**Figure 8:**
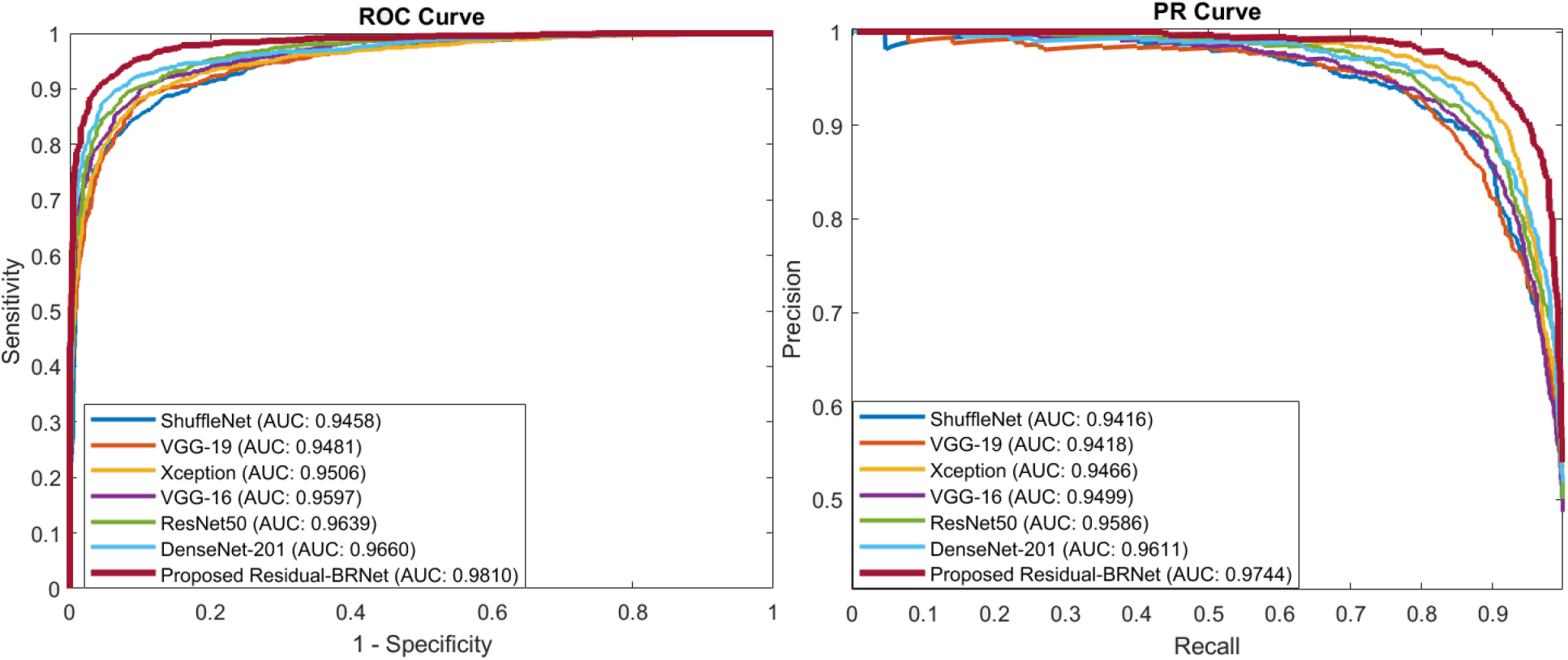
The proposed Residual-BRNet PR and ROC AUC comparison with the existing CNNs

### 5.2 Infectious Regions Analysis

The proposed Residual-BRNet identifies images as COVID-19 infected and channels them to a segmentation CNN for exploring infected regions. Analyzing infected lung lobes is pivotal for understanding infection patterns, and their impact on adjacent lung segments. Additionally, region analysis is essential for quantifying infection severity, potentially assisting in patient grouping and treatment planning for mild versus severe cases. We present PA-RESeg to segment and a series of segmentation CNNs are utilized to measure the model’s learning ability. These models are fine-tuned to detect characteristic COVID-19 imagery features, such as GGO, consolidation, etc. to discriminate typical from infected regions on CT images (Table 3).

#### 5.2.1 Proposed RESeg Segmentation Analysis

The challenge in detecting COVID-19 infection lies in its varied patterns, such as ground-glass opacities, consolidation, and patchy bilateral shadows. Furthermore, the infection’s pattern and extent vary temporally and across individuals, making early-stage differentiation between infected and healthy regions challenging. Thus, a critical aspect is accurately delineating infectious regions with well-defined boundaries within the lungs. To address this, we introduce a novel approach that integrates max- and average pooling within the RESeg and finally, mildly infected region emphasis through PA block. The proposed PA-RESeg demonstrates robust analysis, achieving DSC and IoU of 95.96% and 98.43%, respectively, for lesion regions (Table 3). Notably, precise boundary discrimination is evident from the higher boundary F-score (BFS) value of 98.87%. Comparatively good performing benchmarked against Deeplabv3, PA-RESeg surpasses in DS score, Acc, and IoU. Additionally, generated segmented binary masks illustrate superior visual quality for the proposed PA-RESeg and accurately identify all infected regions (Figure 9). Qualitative analysis confirms the model’s efficacy in segmenting various infection levels (low, medium, high) across different lung lobes, accurately localizing infections whether isolated or multiple distinct segments.

**Figure 9:**
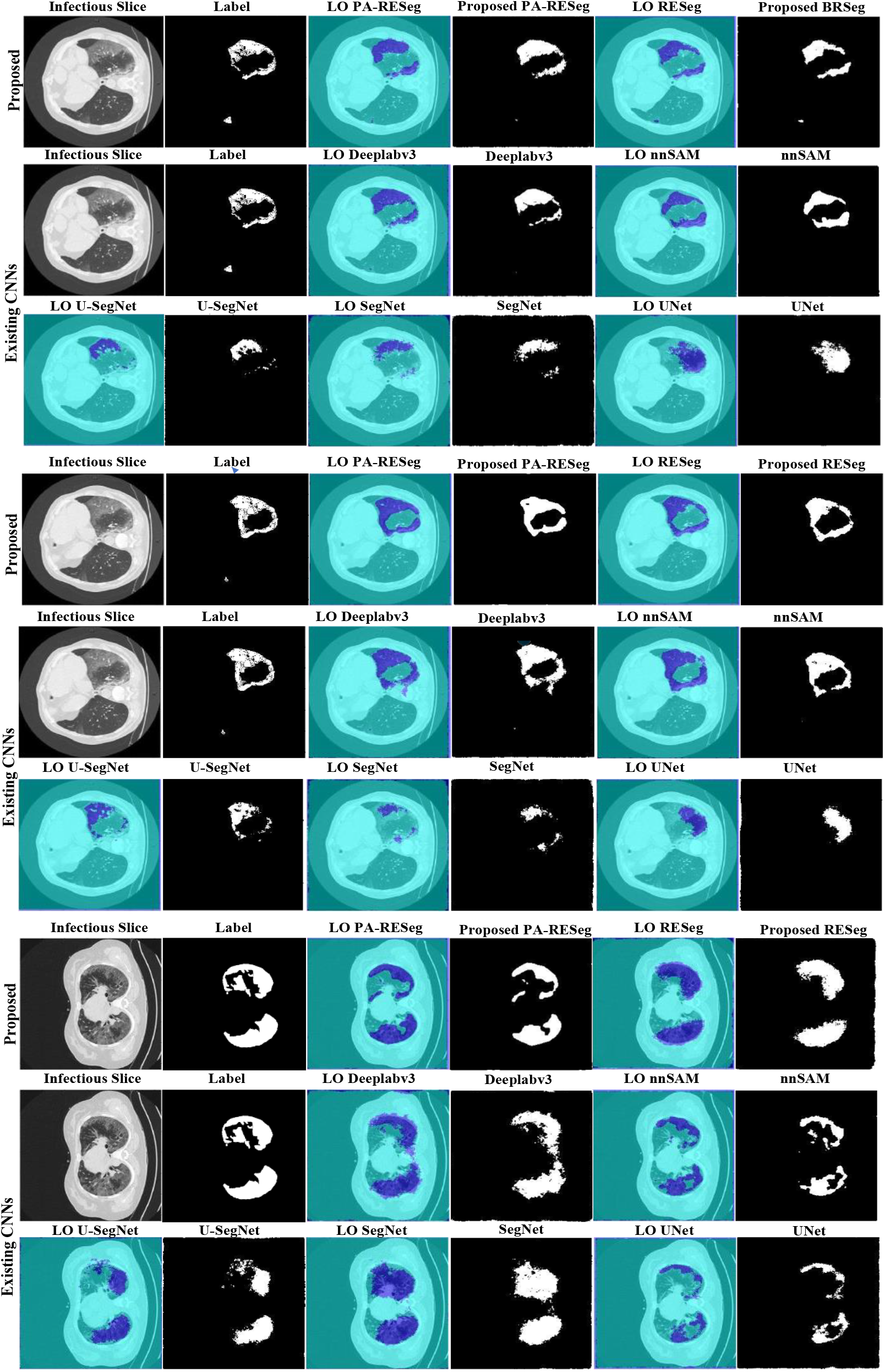
Visual evaluation of the original slice, ground truth, label-overlay (LO), and segmented results of CNNs.

#### 5.2.2 Segmentation Stage Performance Comparison

We evaluated the learning capacity of the proposed PA-RESeg and compared it with current segmentation CNNs (Figure 10 and Table 3). The performance of PA-RESeg is assessed under four metrics: DSC, Acc, IoU, and BFS plots indicating that our proposed outperforms the existing techniques across maximum and minimum scores (Figure 10). The proposed PA-RESeg segmentation model exhibits performance enhancements over existing CNN models on lesion region in BFS (1.34-2.34%), IoU (0.84-2.95%), and DS score (1-3.16%) (Tables 3). Segmented masks generated by PA-RESeg and existing segmentation CNNs are illustrated in Figure 9. Qualitative analysis indicates the consistently good performance of PA-RESeg compared to existing segmentation CNNs. Existing CNNs exhibit poor performance, particularly for mildly infected CT samples, with fluctuations observed in nnUNet, VGG16, and U-Net models, suggesting poor generalization. Among existing models, DeepLabV3 demonstrates good performance, with a DSC: of 95.00%, IoU: of 97.59%, and BFS: of 97.53%. In contrast, our proposed model, though smaller in size, exhibits best performance to highly-capacity DeepLabV3.

**Figure 10:**
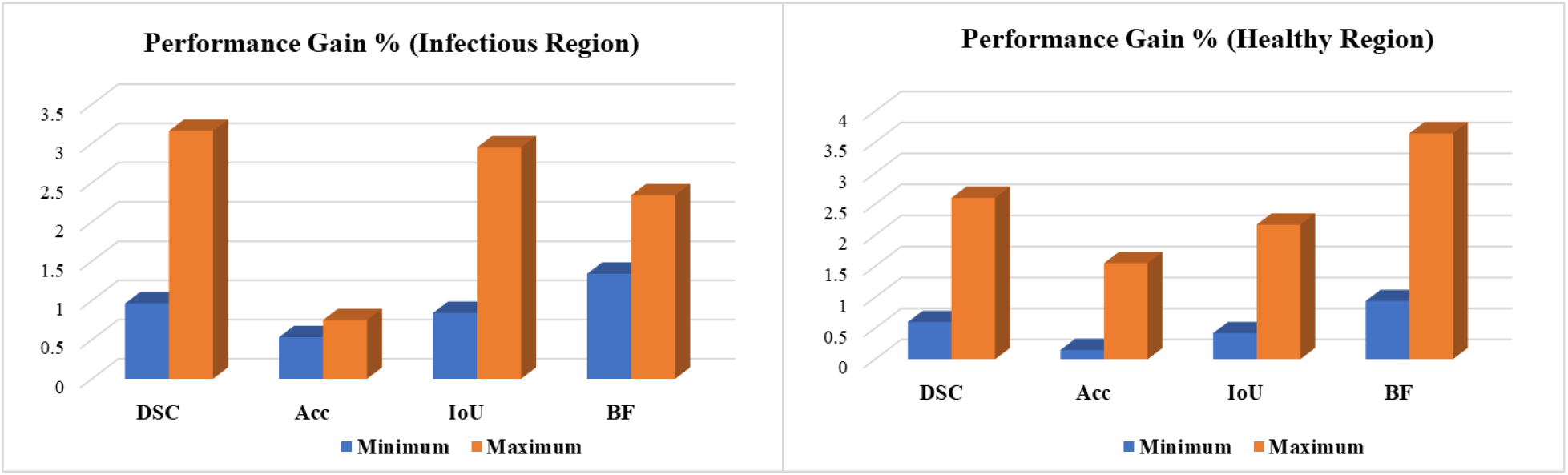
Performance gain of the proposed PA-RESeg over existing segmentation CNNs.

#### 5.2.3 Pixel Attention-Concept

The dataset primarily contains typical healthy lung segments, which can overshadow the COVID-19 infected areas, impacting segmentation model performance. To address this challenge, we implemented an attention concept that integrates pixel weights consistently and enhances segmentation across different infection categories, evident in the visual quality. Notably, there’s a significant enhancement in less severely infected lung sections, with performance gains as shown in Figure 11.

**Figure 11:**
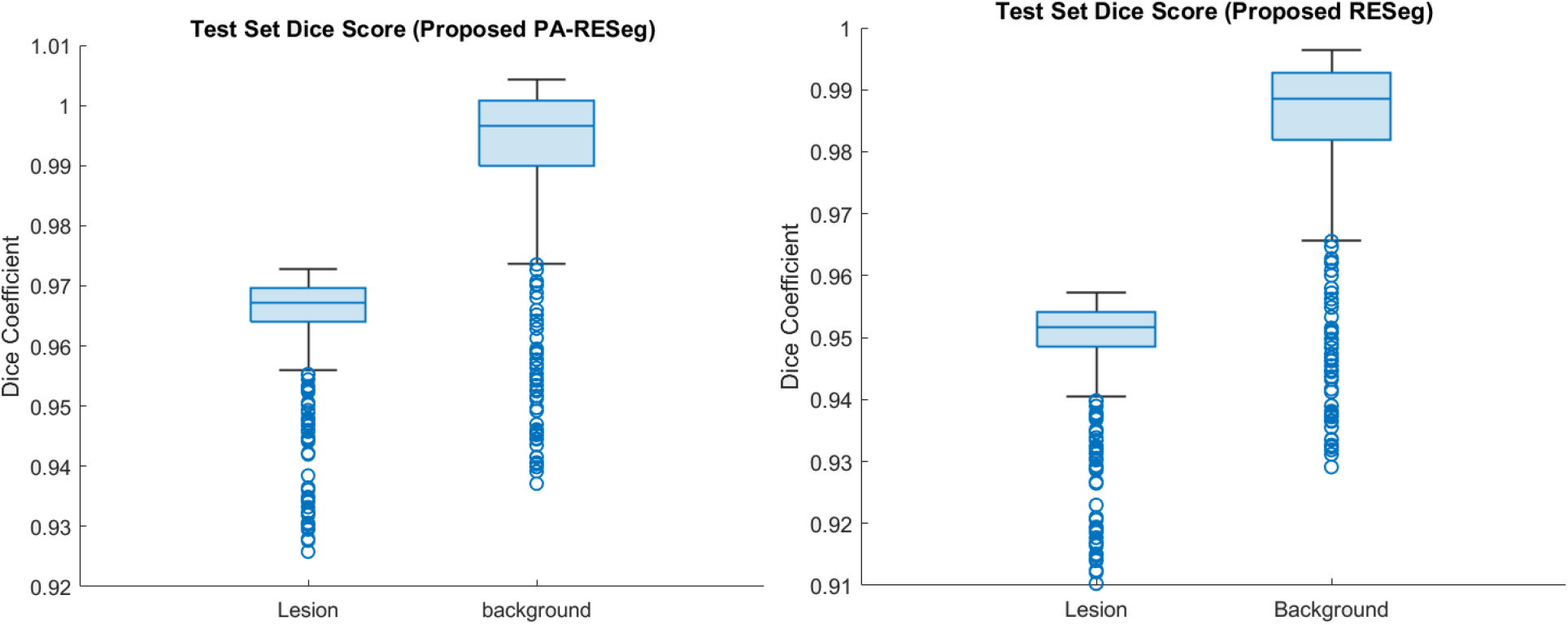
Performance analysis of the proposed segmentation CNNs.

## 6. Conclusions

Prompt identification of COVID-19 infectious patterns is important for efficient prevention and transmission control. A two-stage diagnosis framework is proposed including novel Residual-BRNet CNN for categorizing and new PA-RESeg for scrutinizing COVID-19 infection in CT images. By leveraging various consistent, contrast and texture variations, and structural features, the integrated approach effectively captures COVID-19 radiological patterns. The proposed Residual-BRNet screening CNN demonstrates notable discrimination ability in the initial stage (F-score: 98.01, accuracy: 97.97%, sensitivity: 98.42%) compared to current CNNs, proficiently identifying infectious CT samples. Furthermore, simulations reveal that PA-RESeg achieves precise identification and analysis of infectious images (IoU: 98.43%, DSC: 95.96%). This promising performance validates the efficacy of the two-stage approach in accurately detecting and analyzing COVID-19 infected regions. Such an integrated method aids radiologists in estimating disease severity (mild, medium, severe), whereas single-phase frameworks may lack precision and detailed analysis. Future endeavors will concentrate on applying the proposed framework to larger datasets to enhance real-time diagnostics’ effectiveness and reliability. Additionally, employing dataset augmentation techniques like GANs to generate synthetic examples and expanding the framework by utilizing the integration of CNN with vision transformers to automatically detect infected regions into multi-class patterns will provide comprehensive insights into infectious patterns.

## Acknowledgment

We thank the Artificial Intelligence Lab, Department of Computer Systems Engineering, University of Engineering and Applied Sciences (UEAS), Swat, for providing the necessary resources.

## Institutional Review Board Statement

Not applicable.

## nformed Consent Statement

Not applicable.

## Data Availability Statement

Correspondence and requests for materials should be addressed to Saddam Hussain Khan.

